# ANTI-HUMAN T-LYMPHOTROPIC VIRUS TYPE 1 (HTLV-1) SEROPOSITIVITY IN HAEMATOLOGICAL MALIGNANCIES AT A MAJOR CLINICAL SETTING IN GHANA

**DOI:** 10.64898/2026.07.07.26357496

**Authors:** Frank Awuku, Paul Omoniyi, David Nana Adjei, Makafui Seshie, Kwamena W.C. Sagoe, Amma Anima Benneh-Akwasi Kuma

## Abstract

**Background:** Human T-cell lymphotropic virus - 1 (HTLV-1) is the causative agent of Adult T-cell Leukaemia/Lymphoma (ATLL), a malignancy of CD4+ cells, and HTLV-1-associated Myelopathy/Tropical Spastic Paraparesis (HAM/TSP), a demyelinating disease. Globally, 10–20 million people are infected, though most remain asymptomatic and about 5% progress to severe disease. Transmission occurs mainly through breastfeeding, sexual contact, contaminated needles, and blood transfusion. In Ghana, evidence on the role of HTLV-1 in haematological malignancies remains scarce.

**Methods:** This was a cross-sectional study involving 200 patients with haematological malignancies (Acute Lymphoblastic Leukaemia - 4, Acute Myeloid Leukaemia - 6, Chronic Lymphocytic Leukaemia - 27, Chronic Myeloid Leukaemia - 63, Hodgkin Lymphoma - 21, Multiple Myeloma - 31, Myelodysplasia - 6, Myeloproliferative Neoplasm – 11) at the Haematology Day Care of the Korle-Bu Teaching Hospital. After informed consent was obtained, sera from study participants were tested for anti-HTLV-1 using MP Diagnostics™ GmbH ELISA immunoassay. Data were analysed using R software version 4.0.2 and SPSS version 31.0.0.

**Results:** The study population had a mean age of 49.1±17.7 years, with majority being females (n=109, 54.5%). Of the 200 samples, 16 (8.0 %) were seropositive for HTLV-1, and these were detected in 4 males and 12 females. No statistically significant association was found between HTLV-1 infection and haematological malignancy (exact p = 0.061), sex (p=0.061), and history of blood transfusion (exact p= 1.000).

**Conclusion:** The findings show the seroprevalence of HTLV-1 of 8.0% among patients with haematological malignancies. Although there was no probable association between HTLV-1 and haematological malignancies, screening for HTLV-1 in patients with haematological malignancies may help to unravel the exact contribution in these conditions.

## Introduction

Haematological malignancies are cancers that arise from blood and blood-forming tissues and occur because of genetic alterations in haematopoietic cells in the bone marrow or lymphoid tissue. They include lymphomas, leukaemias, plasma cell myelomas, and myeloproliferative neoplasms ^1,2^. Their aetiology is varied, involving environmental exposures, infections, drugs, radiation, and inherited conditions such as Down’s syndrome ^3^. Approximately 18% of all malignancies including haematologic ones are attributed to infections ^4^. Etiologically, some microorganisms have been linked to the development of malignant lymphomas ^5–7^.

Infections of viral origin are indicated in many haematologic malignancies especially in different subtypes of lymphomas. Notable examples include the Epstein–Barr virus (EBV) which is linked to most Burkitt lymphomas and some histological types of Hodgkin lymphomas ^8^, the Human Immunodeficiency Virus (HIV) which predisposes an individual to lymphomas such as the aggressive B-Cell Non Hodgkin Lymphomas and certain types Hodgkin lymphomas ^9–11^, and Human T-cell Lymphotropic virus type 1 (HTLV-1) which is linked with cases of Adult T-Cell Lymphoma/Leukaemia (ATLL) ^12,13^. A significant association exists between chronic lymphocytic leukemia/small lymphocytic lymphoma (CLL/SLL) and Hepatitis C Virus (HCV) seropositivity in females aged 1–59 years^14^. Other microorganisms like *Helicobacter pylori* are associated with gastric MALT lymphoma ^5,15^.

HTLVs are enveloped RNA viruses of the Retroviridae family discovered in the 1980s ^16^. They have been found to be pathogenic and causally linked to ATLL and HTLV-1-associated Myelopathy/Tropical Spastic Paraparesis (HAM/TSP) ^17–19^. Globally, HTLV-1 infections remain projected at about 20 million people with the highest being in Southern Japan ^20–22^. HTLV-1 is also associated with uveitis and infective dermatitis ^23–27^. Transmission occurs mainly through prolonged breastfeeding, sexual contact, and transfusion of cellular blood products ^28–32^. Studies outside Ghana have detected HTLV-1 in cases of Acute Myeloid Leukaemia (AML), Chronic Myeloid Leukaemia (CML), Acute Lymphoblastic Leukaemia (ALL), Chronic lymphocytic leukaemia (CLL), and lymphomas ^33–37^.

In West Africa, HTLV-1 infections seen among different populations range from 0.5 to 30%. These include school children, commercial sex workers, pregnant women, lepers, neurologic patients, and blood donors ^38–43^. The highest prevalence has been observed in Gabon ^44^. In Ghana, prevalence reached 4.0% in rural and 3.6% in urban populations from 1993 to 2006, increasing with age to 5.9% over 10 years old, with 0.4 – 4.2% (up to 29.49%) among blood donors ^45–48^.

Routine screening for HTLV-1/2 among blood donors has not yet been implemented, exposing recipients of blood or blood products to the risk of HTLV-1/2 transmission, and potentially elevating their susceptibility to lymphomas. Consequently, the role of HTLV-1 in haematological malignancies in Ghana remains unknown. This research focused on the association of HTLV-1 with haematological malignancies at one of the major haematological clinics in Ghana.

## Methods

### Study Site

The study was conducted at the Haematology Department of Korle-Bu Teaching Hospital (KBTH) located in the capital of Ghana, Accra. It is currently Africa’s third-largest hospital, and Ghana’s foremost national referral centre. The hospital has 17 Departments / Units for clinical and diagnostic purposes, and has an estimated daily enrolment of 1,500 patients and about 250 admissions to the hospital. The Department of Haematology is the oldest and largest haematology clinic in country receiving patients from all 16 regions of the country, and other countries especially in the West African subregion. On average the clinic sees 100 new haematological malignant cases annually.

### Study Design and Population

This was a cross-sectional study using simple random sampling to recruit study participants diagnosed with a haematological malignancy at the Haematology Day Care Unit at the Department of Haematology of KBTH. Recruitment commenced on 10 April 2019 and concluded on 30 October 2019. Consented participants included in the study were as follows: Acute Lymphoblastic Leukaemia (ALL) - 4, Acute Myeloid Leukaemia (AML) - 6, Chronic Lymphocytic Leukaemia (CLL) - 27, Chronic Myeloid Leukaemia (CML) - 63, Hodgkin Lymphoma (HD) - 21, Multiple Myeloma (MM) - 31, Myelodysplasia (MDS) - 6, Myeloproliferative Neoplasm (MPN) - 11

### Sample and data Collection

Sample collection was carried out under aseptic conditions. A venous blood sample of 5mls was collected from the antecubital fossa into a clean, plain tube using a vacutainer. The blood was permitted to coagulate completely before centrifugation at 3,000 rpm for 10 minutes using EBA-20 Hettich centrifuge (Andreas Hettich GmbH & Co. KG, Tuttlingen, Germany). Aliquots of the serum were into sterile cryovials and at −80°C prior to analysis. Repeated freezing and thawing were not done. Data abstraction forms were administered to the consenting participants to obtain socio-demographic data, information on type of haematological malignancy, previous history of blood transfusion, date of diagnosis and some laboratory data including haemoglobin levels, haematocrit count, white blood cell count, lymphocyte count, platelet count, liver function test, and lactate dehydrogenase levels.

### Laboratory analysis

Sera from study participants were tested for anti-HTLV I using a commercial enzyme-linked immunosorbent assay (ELISA) (MP Diagnostics™ GmbH, Eschwege, Germany) MP Diagnostics). The testing was done strictly according to manufacturer’s guidelines and instructions. ELISA optical density (OD) measurements were performed using a Spectra II micro-assay plate reader (Molecular Devices, LLC, San Jose, California) at dual wavelength of 450nm-620nm. Cut-offs were calculated as recommended by the manufacturer.^49^

### Data Analysis

Data were recorded in Microsoft Excel file and analysed using R software version 4.0.2 (http://cran.r-project.org) and SPSS version 31.0.0. Descriptive statistics, chi-square tests, and Kruskal-Wallis test were used to analyze the data. A p-value of <0.05 was considered statistically significant.

### Ethical Considerations

This study received ethical approval from the College of Health Sciences Ethical and Protocol Review Committee (EPRC) at the University of Ghana (Protocol ID: CHS-Et/M.S-S.16/2018-2019). Written informed consent was obtained from all participants following a thorough explanation of the study’s objectives, potential risks, and benefits in a language they fully understood, ensuring respect for their autonomy and dignity. All participant data were kept confidential, with identities anonymized through coding using alphanumeric identifiers.

## Results

The outcome of this study suggests some level of endemicity to HTLV-I seropositivity and different reactivity in the different groups of malignancies. Anti-HTLV reactivity was also seen across all malignancies.

### Demographic and Clinical Characteristics

The study included 200 clinically diagnosed patients with haematological malignancy. Females constituted 109 (54.5%) of study participants. The youngest respondent was 16 years and the oldest 92 years old, with a median age of 49.5 ± 17.7 years, but 61 (30.5%) of study participants were 60 years or above. Most of the respondents 125 (62.5%), were employed, 119 (59.5%) were single and majority (81; 40.5%) had attained secondary level education, 73 (36.5%) tertiary level education as indicated in As indicated in **Error! Not a valid bookmark self-reference.** the most prevalent haematological diagnosis was chronic myeloid leukaemia in 63 (31.5%) of the cases. Sixty-eight percent of the patients had no comorbidities, though hypertension alone (19.5%) and diabetes (7.0%) were notable. Disease outcomes showed clinical remission in 148 patients (74.0%) as of study completion, and about a third, 35.5% (71/200), had a history of blood transfusion.

**Table 1.** DEMOGRAPHIC CHARACTERISTICS OF STUDY PARTICIPANTS.

| CHARACTERISTIC | CATEGORY | PROPORTION (%) |
| --- | --- | --- |
| SEX | Male | 91 (45.5%) |
|  | Female | 109 (54.5%) |
| AGE | <20 years | 10 (5.0%) |
|  | 20-29 years | 21 (10.5%) |
|  | 30-39 years | 32 (16.0%) |
|  | 40-49 years | 37 (18.5%) |
|  | 50-59 years | 39 (19.5%) |
| | $\geq 60$ years | 61 (30.5%) |
| OCCUPATION | Employed | 125 (62.5%) |
|  | Unemployed | 11 (5.5%) |
|  | Student | 26 (13.0%) |
|  | Retired | 38 (19.0%) |
| MARITAL STATUS | Single | 119 (59.5%) |
|  | Married | 51 (25.5%) |
|  | Widowed | 30 (15.0%) |
| EDUCATIONAL LEVEL | No Education | 27 (13.5%) |
|  | Primary | 19 (9.5%) |
|  | Secondary | 81 (40.5%) |
|  | Tertiary | 73 (36.5%) |

| CHARACTERISTIC | CATEGORY | PROPORTION (%) |
| --- | --- | --- |
| SEX | Male | 91 (45.5%) |
|  | Female | 109 (54.5%) |
| AGE | <20 years | 10 (5.0%) |
|  | 20-29 years | 21 (10.5%) |
|  | 30-39 years | 32 (16.0%) |
|  | 40-49 years | 37 (18.5%) |
|  | 50-59 years | 39 (19.5%) |
|  | ≥60 years | 61 (30.5%) |
| OCCUPATION | Employed | 125 (62.5%) |
|  | Unemployed | 11 (5.5%) |
|  | Student | 26 (13.0%) |
|  | Retired | 38 (19.0%) |
| MARITAL STATUS | Single | 119 (59.5%) |
|  | Married | 51 (25.5%) |
|  | Widowed | 30 (15.0%) |
| EDUCATIONAL LEVEL | No Education | 27 (13.5%) |
|  | Primary | 19 (9.5%) |
|  | Secondary | 81 (40.5%) |
|  | Tertiary | 73 (36.5%) |

**Table 2.**
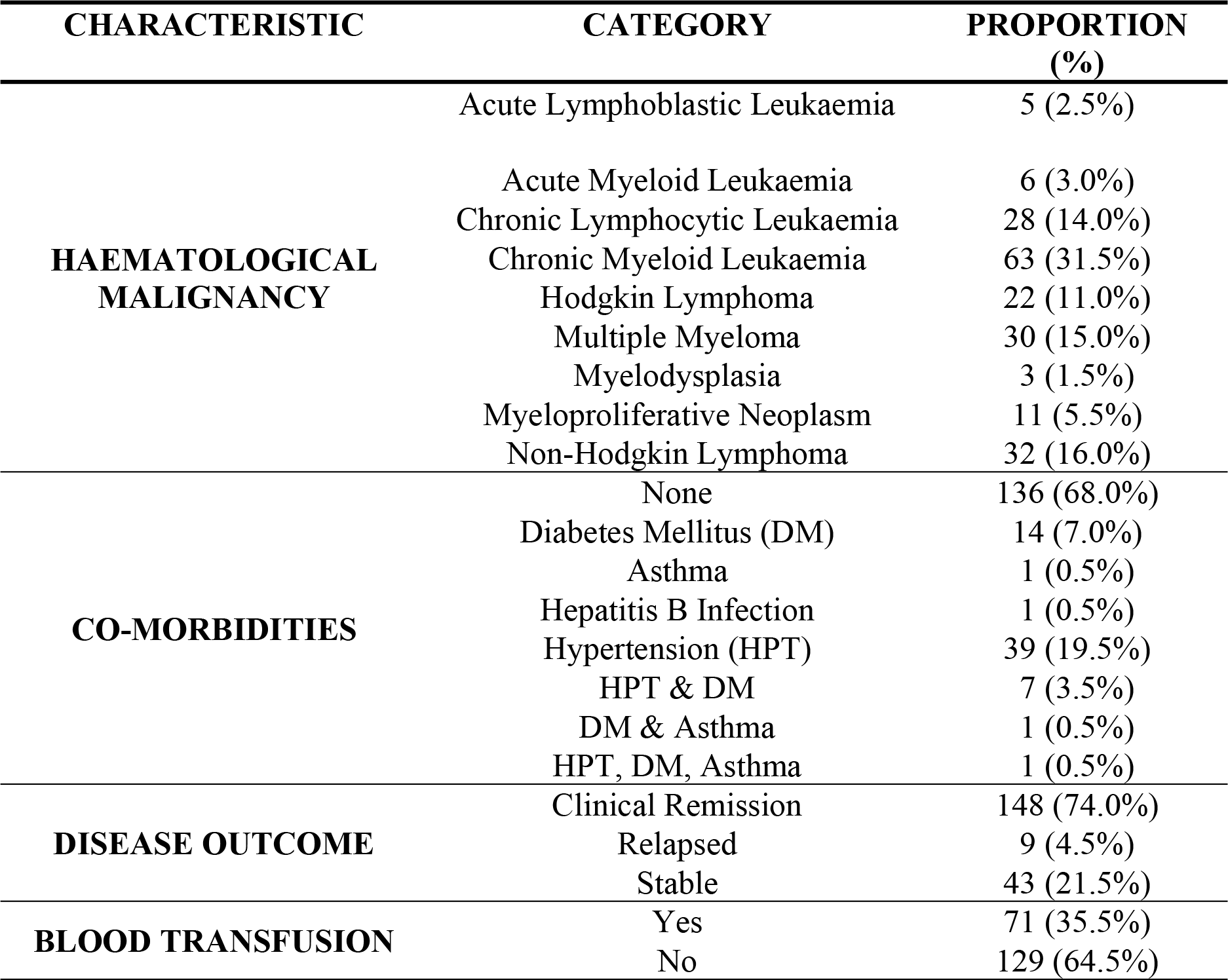
CLINICAL CHARCTERISTICS OF STUDY PARTICIPANTS.

| CHARACTERISTIC | CATEGORY | PROPORTION (%) |
| --- | --- | --- |
| **HAEMATOLOGICAL MALIGNANCY** | Acute Lymphoblastic Leukaemia | 5 (2.5%) |
|  | Acute Myeloid Leukaemia | 6 (3.0%) |
|  | Chronic Lymphocytic Leukaemia | 28 (14.0%) |
|  | Chronic Myeloid Leukaemia | 63 (31.5%) |
|  | Hodgkin Lymphoma | 22 (11.0%) |
|  | Multiple Myeloma | 30 (15.0%) |
|  | Myelodysplasia | 3 (1.5%) |
|  | Myeloproliferative Neoplasm | 11 (5.5%) |
|  | Non-Hodgkin Lymphoma | 32 (16.0%) |
| **CO-MORBIDITIES** | None | 136 (68.0%) |
|  | Diabetes Mellitus (DM) | 14 (7.0%) |
|  | Asthma | 1 (0.5%) |
|  | Hepatitis B Infection | 1 (0.5%) |
|  | Hypertension (HPT) | 39 (19.5%) |
|  | HPT & DM | 7 (3.5%) |
|  | DM & Asthma | 1 (0.5%) |
|  | HPT, DM, Asthma | 1 (0.5%) |
| **DISEASE OUTCOME** | Clinical Remission | 148 (74.0%) |
|  | Relapsed | 9 (4.5%) |
|  | Stable | 43 (21.5%) |
| **BLOOD TRANSFUSION** | Yes | 71 (35.5%) |
|  | No | 129 (64.5%) |

Laboratory parameters revealed a high prevalence of abnormalities among study participants. (**Table 3**) Anaemia (haemoglobin (HB) <12 g/dL in women, <13 g/dL in men) affected 142 (71.0%) patients, while haematocrit (HCT) abnormalities were more common in 139 (69.5%) patients. Leukocytosis or leukopenia (abnormal Total White Blood Cell count (TWBC) occurred in 67 (33.5%) patients, with lymphopenia (abnormal lymphocyte count (LYM)) less frequent in 25 (12.5%) patients, and thrombocytopenia (low platelet count (PLT) in 41 (20.5%) patients. Disease-related markers showed elevated lactate dehydrogenase (LDH) in 131 patients (65.5%), a strong indicator of tumour burden, and liver function test (LFT) abnormalities in 60 (32.0%) patients. These patterns reflect advanced disease and systemic involvement typical in lymphoma cohorts.

**Table 3.** HAEMATOLOGICAL PARAMETERS OF STUDY PARTICIPANTS.

| PARAMETER | ABNORMAL, N (%) | NORMAL, N (%) |
| --- | --- | --- |
| HB g/dl | 142 (71.0%) | 58 (29.0) |
| HCT % | 139 (69.5%) | 61 (30.5%) |
| TWBC x10<sup>9</sup>/L | 67 (33.5%) | 133 (66.5%) |
| LYM x10<sup>9</sup>/L | 25 (12.5%) | 175 (87.5%) |
| PLT x10<sup>9</sup>/L | 41 (20.5%) | 159 (79.5%) |
| LDH | 131 (65.5%) | 69 (34.5%) |
| LFT | 64 (32.0%) | 136 (68%) |

### Seroprevalence of HTLV-1

Of the 200 participants, 16 (8.0%) tested seropositive for HTLV-1 antibodies. This prevalence was higher in females 12 (11%) than in males 4 (4.4%) but this was not statistically significant (exact p = 0.086). The highest prevalence was observed among the 30-39 age group (5, 15.6%), but there was no correlation between age and HTLV −1 infection (exact p = 0.326).

All cases confirmed seropositivity with OD ratios >1.0 (median 10.56, range 1.098–14.110). Myeloid malignancies (CML, MDS) exhibited highest reactivity (ratios 5.153–14.110), while Non-Hodgkin Lymphoma showed greatest variability (1.972–12.298) (***Figure 1***). There were no statistically significant differences for optical differences between any of the groups (p = 0.5).

**Figure 1.**
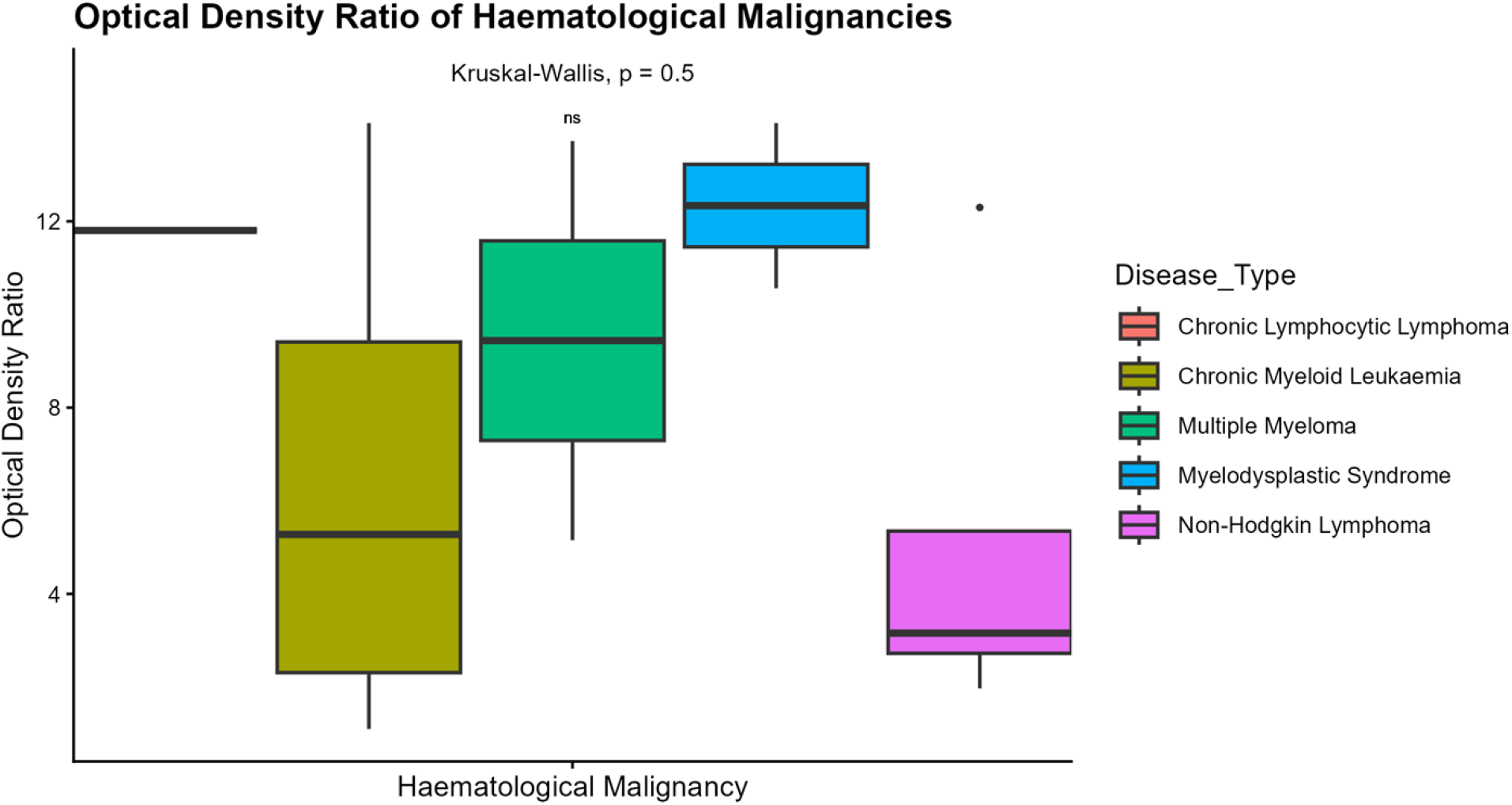
OPTICAL DENSITY RATIO OF HTLV-1 CASES.

### Associations of Clinical Characteristics and HTLV-1 Infection

Of the 16 (8.0%) study participants seropositive for HTLV-1 antibodies, 7 (43.75%) were diagnosed with Non-Hodgkin Lymphoma, 5 (31.25%) had Chronic Myeloid Leukaemia, 2 (12.5%) had Multiple Myeloma, and one each (6.25%) had Chronic Lymphocytic Leukaemia and Myelodysplasia. But there was a tendency towards some malignancies having anti-HTLV 1 seropositivity (exact p=0.061). No significant associations were found between HTLV-1 infection and blood transfusion history (exact p=1.000). Similarly, no significant differences were observed in haematological parameters such as haemoglobin, total white blood cells, lymphocytes, or platelets between HTLV-1-seropositive and HTLV-1-seronegative participants.

### Treatment Outcomes

The majority of HTLV-1 seropositive participants (number, 75.0%) were in clinical remission at the time of the study. No significant association was found between HTLV-1 seropositivity and treatment outcomes (p=0.820).

## Discussion

HTLV-1 is transmitted through cell-to-cell contact, most commonly via blood transfusion, sexual contact, and breastfeeding ^13^, and has been associated with diseases that are rare and quickly fatal, constituting real challenges in determining the prevalence of induced pathologies and/or impact on public health ^50,51^. Understanding its prevalence in different populations is critical, as such information guides prophylactic measures to reduce transmission. HTLV-1 infection is chronic, untreatable, and a recognized risk factor for haematologic malignancies, particularly ATLL^13^.

This study investigated HTLV-1 seropositivity in patients with haematological malignancies at Korle-Bu Teaching Hospital and found a seroprevalence of 8.0% (16/200). More women were affected (54.5%), consistent with local and international data on leukaemia patients ^52,53^. Compared with other studies, the prevalence we observed is lower than values from Dominica (38.6%) and Iran (30%) but aligns with findings from Nigeria (5.1%) and is higher than rates reported among Ghanaian blood donors (3.3%) and pregnant women (2.1%), and 7.3% among individuals attending the main teaching hospital in Gabon ^38,44,46,54–56^. The higher prevalence in this cohort likely reflects the elevated risk among patients with malignant haematological disease. Although no significant association between HTLV-1 seropositivity and haematological malignancies was found, this could be due to the small number of patients sampled. We also noticed a high prevalence (50%; 7/14) of HTLV-1 among participants with Non-Hodgkin Lymphoma, similar to the 45% prevalence reported in a group of Iraqi patients with Non-Hodgkin lymphoma^57^, and higher than 18.8% (10/54) prevalence in an Iranian population^58^.

Although no significant association was observed between transfusion history and HTLV-1 seropositivity, a higher proportion of seropositive patients had received transfusions (8.5%) compared with those without transfusion history (6.2%). While not statistically significant, this trend has been reported in other studies where transfusion, particularly repeated transfusions, was a major risk factor ^56^. Age-specific analysis showed no cases under 30 years, with the highest prevalence (12.5%) occurring in the 30–39 years age group, consistent with other studies linking HTLV-1 seropositivity with increasing age ^59^. Although there is a higher risk of HTLV-1 infection with multiple sexual partners, married participants in this study had the highest prevalence.

Most study participants were in clinical remission, and treatment outcomes were not associated with HTLV-1 seropositivity. Among HTLV-1 seropositive cases, 78.6% were in clinical remission at the time of the study, suggesting effective control of malignancy under current treatment regimens. No significant associations were found between HTLV-1 status and haematological parameters (Hb, WBC, lymphocytes, platelets), which may reflect the stable clinical state of most patients at the time of recruitment. Similar observations were made elsewhere ^60^, where it was reported that differences in haematological indices are more apparent at diagnosis.

## Conclusion

To our knowledge, this study provides the first evidence of a relatively high prevalence of HTLV-1 among patients with haematological malignancies in Ghana, much higher than in general population cohorts. The findings support a probable role for HTLV-1 in the aetiology of blood cancers and highlight the need for routine HTLV-1 screening and preventive strategies in Ghana.

The results of this study have shown the seroprevalence of HTLV-1 of 8.0% among patients with haematological malignancies, and probable association between some haematological malignancies and HTLV-1. Consequently, it is possible to consider HTLV-1 infection as a risk factor for other haematological malignancies other than lymphoid malignancies. There was no significant association of HTLV-1 with previous history of blood transfusion as well as some haematologic parameters (Hb, lymphocyte count, total WBC and platelet count). This study revealed that the seroprevalence of HTLV-1 in hematologic malignancies is higher compared to the seroprevalence of the general healthy population reported in Ghana. Further larger prospective studies are needed to corroborate the current evidence.

### Limitations and Recommendations

This study had some limitations. Firstly, the minimal sample size may not clearly represent the HTLV-1 burden in the country. Secondly, genomic analysis was not conducted on positive cases to quantify viral load and identify the circulating genotypes. Lastly, due to its cross-sectional nature, HTLV-1 positive participants were not continuously followed to record and analyse clinical status.

Nonetheless, we recommend prospective and multi-centre research on the seroprevalence of HTLV-1 seropositivity among blood donors, and other at-risk. A standard and simultaneous national epidemiological survey should be carried out in all the geopolitical zones of Ghana to establish the true seroprevalence of HTLV-1 infection as an initial step in determining its impact on haematological diseases.

## Acknowledgments

We thank the staff and patients of the Haematology Department, Korle-Bu Teaching Hospital, for their support and participation in this study. We also acknowledge the Department of Medical Microbiology, University of Ghana, for their technical assistance.

## Data Availability

All data has been made available in the paper.

